# Fully Automated Deep Learning-Based Pipeline for Evans Index Measurement from Raw 3D MRI

**DOI:** 10.64898/2025.11.30.25341302

**Authors:** Siavash Shirzadeh Barough, Murat Bilgel, Ameya Moghekar, Catalina Ventura, Mark G. Luciano, Abhay Moghekar

## Abstract

Ventriculomegaly is a key neuroimaging feature in conditions such as normal pressure hydrocephalus (NPH) and other disorders of cerebrospinal fluid (CSF) dynamics. The Evans Index (EI), defined as the ratio of the maximal frontal horn width to the maximal inner skull diameter, remains a simple and widely used marker for ventricular enlargement. However, manual EI measurement is subject to observer variability and dependent on proper alignment to the anterior commissure-posterior commissure (AC-PC) plane, limiting reproducibility in large-scale and multi-center studies.

We present a fully automated deep learning-based pipeline for EI calculation directly from raw T1-weighted MPRAGE MRI scans. The pipeline integrates (i) landmark detection using the BrainSignsNet model, (ii) rigid AC-PC alignment, and (iii) robust segmentation of the lateral ventricles (LV) and intracranial volume (ICV) via nnU-Net models, including a custom ventricular network trained on 1,300 annotated scans enriched for hydrocephalus. The Evans Index is then derived from automated measurement of frontal horn width and inner skull diameter at the aligned axial slice.

Internal validation using data from the Baltimore Longitudinal Study of Aging, BIOCARD, and Johns Hopkins cohorts demonstrated high segmentation accuracy (Dice coefficient = 0.98). External validation on the PENS trial, including pre- and post-shunt NPH scans, showed excellent agreement with expert manual EI measurements (mean bias = 0.0068, mean absolute error = 0.0103, r = 0.96, p < 0.001). Bias analyses revealed no significant association between measurement error and age, sex, or ventricular volume

This fully automated, orientation-standardized method achieves accurate and reproducible Evans Index measurement across diverse MRI datasets. By eliminating manual intervention, the pipeline enhances scalability for large neuroimaging cohorts and provides a reliable tool for clinical assessment, screening and monitoring of ventriculomegaly, especially in NPH.

## Introduction

Ventriculomegaly is a key neuroimaging finding in several neurological conditions, including normal pressure hydrocephalus (NPH)^1,2^, neurodegenerative diseases^3^, and other disorders affecting cerebrospinal fluid (CSF) dynamics^4,5^. Accurate quantification of ventricular enlargement is essential for diagnosis, disease monitoring, and surgical planning, particularly in NPH where intervention can substantially improve patient outcomes. The Evans Index (EI), defined as the ratio of the maximal frontal horn width to the maximal internal diameter of the skull at the same axial level, is one of the most widely used, simple, and reproducible measures of ventricular size in clinical practice.^6^ Despite its simplicity, EI remains a critical diagnostic criterion in consensus guidelines, often used as a threshold marker to distinguish pathological from age-related ventricular dilation.^7^

Traditionally, EI measurement is performed manually by trained radiologists or neurologists, typically on two-dimensional slices derived from MRI or CT scans. While conceptually straightforward, manual EI calculation is time-consuming, subject to inter- and intra-observer variability, and dependent on the selection of an optimal slice aligned with the anterior commissure–posterior commissure (AC–PC) plane ^8^. Variations in head orientation, image quality, and slice selection criteria can lead to inconsistent results, particularly in multi-center studies or large-scale clinical trials. These challenges highlight the need for standardized, reproducible, and efficient approaches to EI measurement.

The advent of high-resolution three-dimensional MRI sequences, such as T1-weighted MPRAGE, combined with recent advances in artificial intelligence (AI) and deep learning, offers new opportunities to automate morphometric analyses with high precision. AI-driven methods can leverage the full volumetric dataset to identify anatomical landmarks, correct for head rotation, and consistently determine the optimal measurement plane. Automated pipelines can thus minimize human bias, improve reproducibility, and significantly reduce processing time, enabling integration into both research workflows and routine clinical practice. However, despite the promise of AI, few studies have specifically addressed fully automated EI measurement in unprocessed T1 MPRAGE scans, especially in a manner robust to head positioning and inter-scan variability.

In this study, we develop and validate a fully automated, AI-based method for EI calculation directly from raw T1 MPRAGE MRI scans. Our pipeline integrates deep learning-based landmark detection, automated AC–PC alignment, and robust ventricular segmentation to ensure accurate and reproducible EI measurements across a wide range of scan orientations and acquisition parameters. We evaluate the method’s performance against expert manual measurements and assess its robustness across varying head positions. By eliminating manual intervention, this approach has the potential to standardize EI quantification, facilitate large-scale neuroimaging studies, and enhance clinical decision-making in disorders characterized by ventricular enlargement.

## Method

### Data Acquisition

T1-weighted MRI data for pretraining, training, and internal validation were drawn from the BLSA^9^ and the Johns Hopkins Hydrocephalus Clinic. BLSA scans comprised: at 1.5 T, SPGR acquired on three GE Signa systems (0.94 × 0.94 × 1.5 mm voxels; flip angle 45°; TE = 5 ms; TR = 35 ms; TI = 0 ms) plus one Philips Intera 1.5 T SPGR system with identical parameters; and at 3 T, sagittal MPRAGE acquired on three Philips Achieva systems (1 × 1 × 1.2 mm voxels; flip angle 8°; TE = 3.2 ms; TR = 6.5–6.8 ms; TI = 0 ms). Johns Hopkins Clinic scans were obtained on a Siemens Verio 3 T (0.78 × 0.78 × 0.80 mm voxels; flip angle 9°; TE = 3.13 ms; TR = 1800 ms; TI = 900 ms). Additionally, for the Placebo-controlled Efficacy of iNPH Shunting (PENS)^10^ study we acquired multi-echo T1-weighted MPRAGE with the same protocol across all sites (scan time = 2 min 54 s; resolution = 1 × 1 × 1 mm^3^; FOV = 240 × 240 × 180 mm^3^; 180 slices; TR/TE/TI = 14/2.1/900 ms; flip angle = 9°).

### Data preprocessing

Prior to training, validation, and inference of the method described in this study, all MRI data underwent a standardized preprocessing pipeline to ensure consistency and reliability. The pipeline included N4 bias field correction to mitigate intensity inhomogeneities^11^. Subsequently, rigid registration to the MNI152 standard space was performed to align images to a common anatomical template, correcting for differences in head orientation and facilitating inter-subject comparisons. Finally, all the scans were then reoriented to Right Anterior Superior (RAS) and then using the AC and PC coordinates extracted via BrainSignsNet^12^ aligned in sagittal and axial axes according to the original method of EI measurement.

### Data annotations

All available scans were first pre-processed and subjected to automated ventricular segmentation using the Deep Learning–based MUlti-SEquence framework (DLMUSE).^13^ Following this initial pass, segmentations were refined using the Medical Segment Anything (Medical SAM) approach, which leveraged interactive point-based prompts to correct residual boundary errors and ensure anatomical fidelity.^14^

Evans Index was measured in all the images of external validation dataset manually, adhering to the classical measurement protocol.^7^ First, the anterior commissure (AC) and posterior commissure (PC) were localized to establish a consistent axial reference plane. On slices parallel to this reference plane, the maximal distance between the frontal horns of the lateral ventricles, defined as the ventricular portions anterior to the interventricular foramen (IVF), was measured. The maximal inner skull diameter was then determined on the same axial slice. The Evans Index was calculated as the ratio of the frontal horn width to the inner skull diameter.

### Segmentation models

Inner skull segmentation was performed using the pretrained Deep Learning-based Inner Cranial Volume (DLICV) model, which has been validated for robust delineation of the inner table of the cranial vault across diverse MRI protocols (https://github.com/CBICA/DLICV).

Lateral ventricle segmentation was achieved by training a custom 3D nnU-Net v2^15^ model on an annotated dataset specifically enriched for marked ventricular dilation, reflective of our target pathology. We curated 1,622 T1-weighted brain MRI scans with manual lateral-ventricle segmentation masks and split these into an 80 % training set (n = 1,300) and a 20 % internal validation set (n = 322). Training followed the default nnU-Net v2 pipeline configuration, with one critical modification: flipping was disabled in the augmentation module to preserve ventricular laterality and prevent misclassification of left versus right ventricles. Model checkpoints were evaluated on the internal validation subset using the Dice similarity coefficient (DSC), and the highest-performing model was selected for further testing. Generalizability was assessed on an external cohort comprising patients with normal-pressure hydrocephalus (NPH), imaged both before and after shunt surgery, to confirm robustness of ventricle segmentation across clinical presentations.

After segmentation tasks, postprocessing steps were then applied to refine the segmentation output. Specifically, small spurious clusters disconnected from the primary intracranial volume were removed, and topological corrections were performed to fill internal voids within the main mask, thereby ensuring a continuous intracranial volume.

### Landmark Detection and Scan Alignment

We employed the BrainSignsNet model, previously trained and externally validated on a large set of scans manually annotated for the anterior commissure (AC), posterior commissure (PC), and interventricular foramen (IVF). In evaluations, the model achieved a mean absolute error (MAE) of 1.82 mm (standard deviation: 1.55 mm) in the distance between predicted and manually annotated landmarks. For pitch correction, the model demonstrated an average MAE of 1.57° (standard deviation: 1.44°).

### Training and Validation Setup

All model training and inference were performed on a high-performance workstation equipped with two NVIDIA RTX 4090 GPUs, 128 GB of RAM, and an AMD Threadripper Pro WRX80 CPU. All training sessions ran until convergence.

### Evans Index Calculation

To automatically calculate the Evans Index from brain MRI scans, we implement a structured multi-step pipeline (Figure 1). Step 1: MRI scans are loaded in NIfTI format (or converted from DICOM if required), reoriented to canonical RAS orientation, and pre-processed using N4 bias field correction and rigid registration to MNI space to ensure consistency across subjects. Step 2: Key anatomical landmarks, the anterior commissure (AC), posterior commissure (PC), and interventricular foramen (IVF), are localized using the BrainSignsNet model. Step 3: Segmentation of the lateral ventricles (LV) and intracranial volume (ICV) is performed using nnU-Net models. Step 4: Brain volumes and masks are rotated and realigned based on the AC–PC coordinates, such that the AC–PC line becomes horizontal in the sagittal plane and vertical in the axial plane. The frontal horns of the lateral ventricles are then identified using the IVF as a reference point. Step 5: The maximum distance between the two ventricles, measured along a line perpendicular to the sagittal plane, is obtained. Step 6: At the corresponding axial slice (same Z-coordinate) of the ICV mask, the maximum inner skull width is measured. Step 7: The Evans Index is computed as the ratio of the frontal horn width to the maximum inner skull diameter.

**Figure 1.**
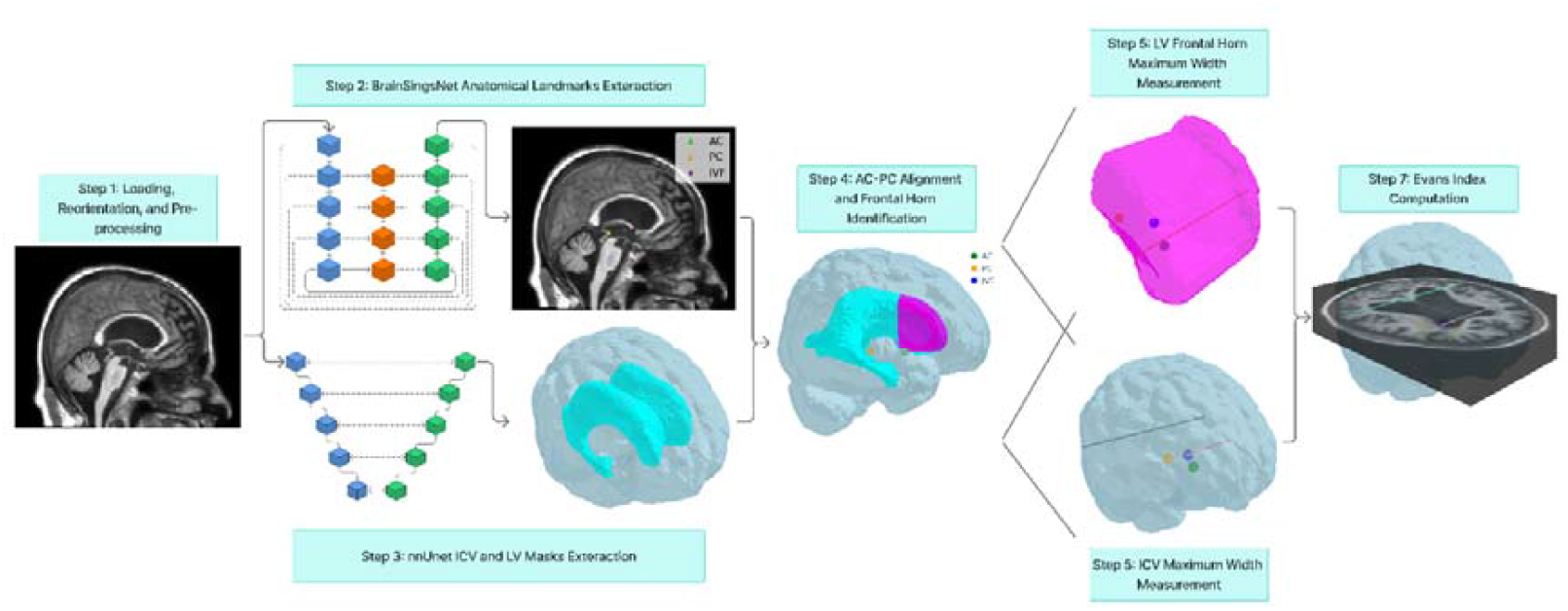
Automated Evans Index calculation pipeline. AC: anterior commissure; PC: posterior commissure; IVF: interventricular foramen (foramen of Monro); ICV: intracranial volume; LV: lateral ventricles.

### Bias Assessment

Potential systematic biases in automated Evans Index measurements were evaluated by examining their relationship with demographic variables (age and sex) and anatomical characteristics (total lateral ventricular volume). Additionally, the impact of rotational misalignment relative to the AC-PC line was assessed by applying controlled rotations around three orthogonal axes (pitch, yaw, and roll) at eight predefined angles (±5°, ±10°, ±15°, ±20°). Measurement errors were analyzed to determine whether discrepancies were influenced by patient characteristics, baseline ventricular size, or deviations from the reference plane during image acquisition.

### Statistical analysis

Continuous variables are reported as mean ± standard deviation, and categorical variables as count (percentage). Normality was assessed visually and with the Shapiro–Wilk test where applicable. Between-group comparisons for normally distributed continuous variables were performed using independent two-sample t-tests, while comparisons involving more than two groups were conducted using analysis of covariance (ANCOVA) with age and sex included as covariates. Pearson’s correlation coefficient (r) was used to quantify associations between continuous variables. Agreement between automated and manual Evans Index measurements was assessed using Bland–Altman analysis, reporting mean bias, standard deviation, and 95% limits of agreement, along with the mean absolute error. The Wilcoxon signed-rank test was applied to evaluate the effect of rotational perturbations on measurement error. All statistical tests were two-tailed, with p < 0.05 considered statistically significant. Landmark detection accuracy was expressed as the mean absolute Euclidean distance (mm) between predicted and manually annotated commissure coordinates. Segmentation accuracy was quantified using the three-dimensional Dice similarity coefficient.

## Results

### Demographics

MRI scans were obtained from four cohorts (Table 1). The Baltimore Longitudinal Study of Aging (BLSA) contributed 1,032 scans from 1,032 participants (mean age, 73.32 ± 13.36 years; 45.6% male, 54.4% female) and the BIOCARD study contributed 395 scans from 395 participants (mean age, 64.22 ± 11.02 years; 40.8% male, 59.2% female). The Johns Hopkins Hydrocephalus Clinic provided 195 scans from 195 patients (mean age, 71.54 ± 8.30 years; 53.8% male, 46.2% female). Data from these three cohorts were used for model training and internal validation. The PENS cohort contributed 190 scans from 99 participants (mean age, 75.00 ± 5.70 years; 51.5% male, 48.5% female) and served as the external validation dataset.

**Table 1.** Demographic characteristics of participants in the training and validation datasets. Age is reported as mean ± standard deviation, with distributions further stratified by sex and age group.

| Cohort | Number of samples | Number of Participants | Age | Sex | Use |
| --- | --- | --- | --- | --- | --- |
| BLSA | 1,032 | 1,032 | 73.32 (13.36) | Male = 471 (45.63%)<br>Female = 561 (54.36%) | Training and Internal validation |
| BIOCARD | 395 | 395 | 64.22 (11.02) | Male = 161 (40.75%)<br>Female = 234 (59.24%) | Training and Internal validation |
| Johns Hopkins Hydrocephalus Clinic | 195 | 195 | 71.54 (8.30) | Male = 105 (53.8%)<br>Female = 90 (46.1%) | Training and Internal validation |
| PENS | 190 | 99 | 75.00 (5.70) | Male = 51 (51.51%)<br>Female = 48 (48.48%) | External Validation and Evans Index validation |

### Segmentation Models

After training the nnU-Net V2 model for nearly 1,000 epochs (until convergence) on MRI data from the Baltimore Longitudinal Study of Aging (BLSA), BIOCARD, and Johns Hopkins cohorts, it achieved a Dice similarity coefficient of 0.98 during internal validation. When this model was applied to all scans from the PENS trial for external validation, it sustained its performance, yielding an overall Dice score of 0.98.

### Evans Index Measurement Performance

In pre-shunt scans (Figure 2a), our automated Evans Index measurements, derived from our trained ventricular segmentation network, demonstrated a mean bias of 0.0053 (SD 0.0101) and 95% limits of agreement from –0.0146 to +0.0251; post-shunt scans (Figure 2b) yielded a mean bias of 0.0083 (SD 0.0120) with limits of agreement between –0.0153 and +0.0318. Across all scans, the combined bias was 0.0068 (SD 0.0112; limits –0.0152 to +0.0287), with a mean absolute error of 0.0103 and a strong correlation to manual Evans Index measurements (r = 0.9585, p < 0.001) (Figure 2c). By comparison, the DLMUSE-based segmentation approach produced a pre-shunt bias of 0.0101 (SD 0.0094; limits –0.0083 to +0.0285) and a post-shunt bias of 0.0121 (SD 0.0111; limits –0.0096 to +0.0338). Overall, DLMUSE exhibited a combined bias of 0.0110 (SD 0.0102; limits –0.0090 to +0.0311), a mean absolute error of 0.0116, and a significant correlation with manual measurements (r = 0.9659, p < 0.001) (Supplementary Figure 2). These findings indicate that both segmentation methods yield highly accurate and reproducible Evans Index estimations, with our trained model achieving lower systematic error.

**Figure 2.**
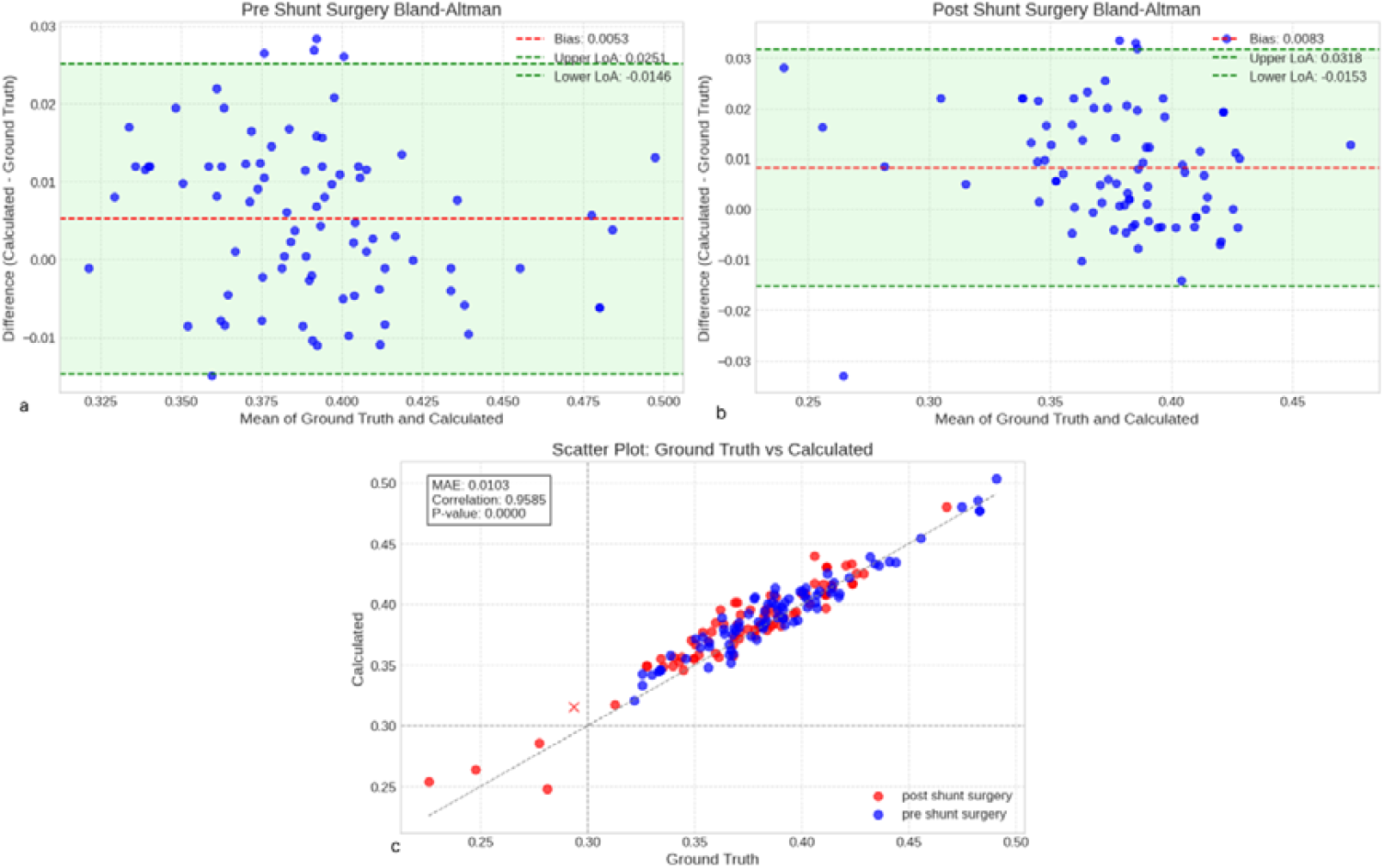
Performance analysis of automated Evans Index calculation. (a–b) Bland–Altman plots comparing automated Evans Index measurements with expert annotations for pre-shunt (a) and post-shunt (b) scans. (c) Scatter plot showing correlation between automated and expert measurements, with pre-shunt scans in blue and post-shunt scans in red. Point(s) marked with X represent misclassified data at a 0.30 threshold.

### Error and Bias Analysis

To evaluate potential systematic biases in the automated Evans Index measurements, we first examined the relationship between measurement error and demographic or anatomical variables. Across the study cohort, absolute Evans Index errors showed no significant correlation with age, sex, or total lateral ventricular volume (all p > 0.05), indicating that the observed discrepancies were not driven by these factors. This suggests that the automated method is not inherently biased by patient demographics or baseline ventricular size (Figure 3).

**Figure 3.**
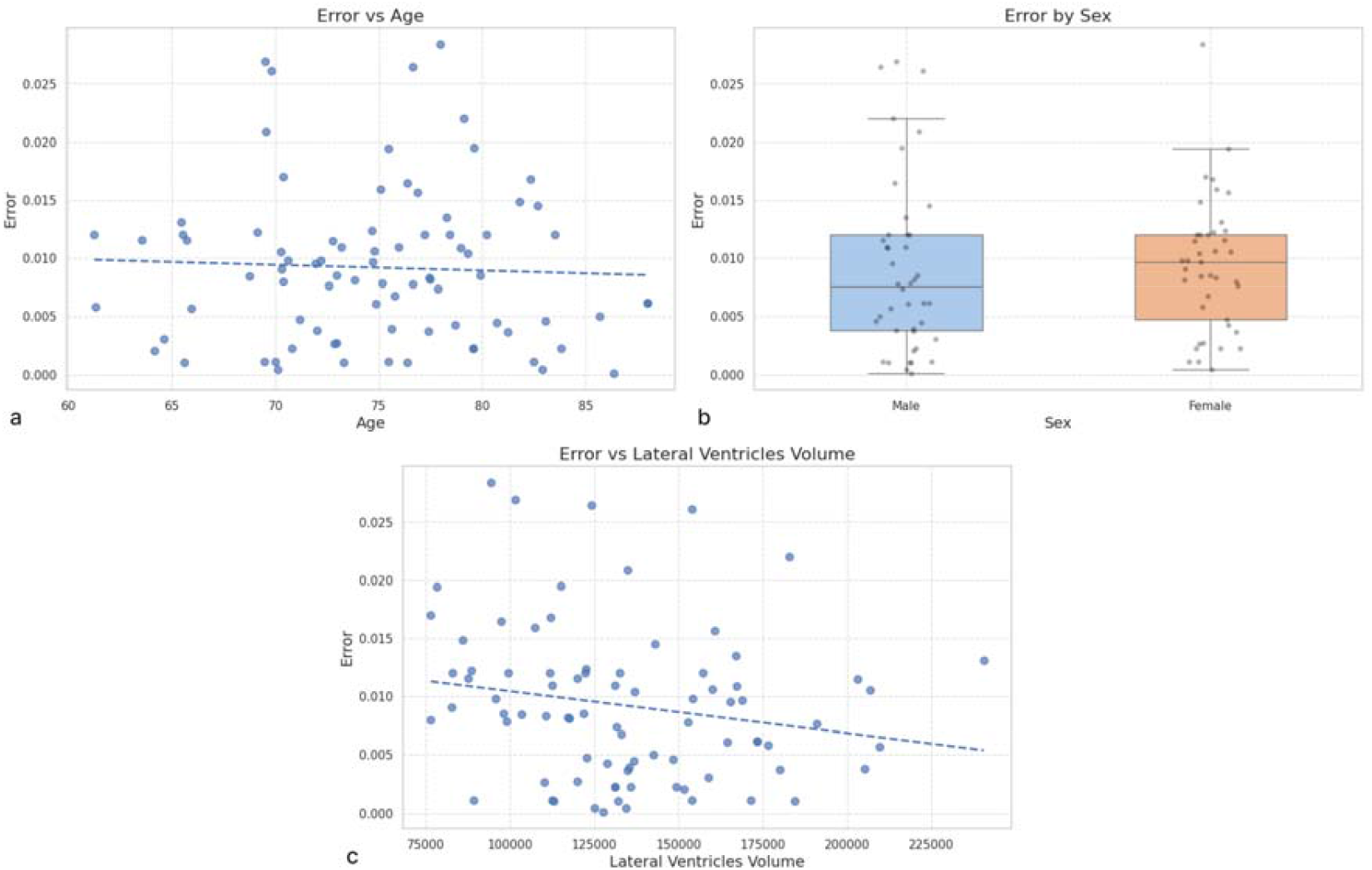
Analysis of potential systematic biases in automated Evans Index measurements. (a) Scatter plot of absolute Evans Index error versus age. (b) Boxplot of absolute error stratified by sex. (c) Scatter plot of absolute error versus total lateral ventricular volume.

The impact of rotational misalignment relative to the PC–AC line was then systematically quantified across three orthogonal axes (pitch, yaw, and roll) (Supplementary Figure 2), each evaluated at eight rotation angles (±5°, ±10°, ±15°, ±20°) (Figure 4). Wilcoxon signed-rank tests revealed statistically significant increases in absolute measurement error for all rotation angles compared with the neutral orientation (all p < 0.001). The magnitude of error was symmetric for positive and negative rotations of the same degree, with maximal Wilcoxon statistics observed at ±20° for each axis. Notably, yaw and roll rotations produced the largest rank-sum values at extreme angles, reflecting particularly pronounced deviations in Evans Index measurements under large axial rotations. Even small deviations (±5°) produced highly significant differences (all p < 0.0001), underscoring the sensitivity of the Evans Index to subtle misalignments. These findings highlight the necessity of maintaining consistent head positioning relative to the AC–PC plane during acquisition, as rotational variance—especially beyond ±10°—introduces systematic, morphology-independent measurement errors.

**Figure 4.**
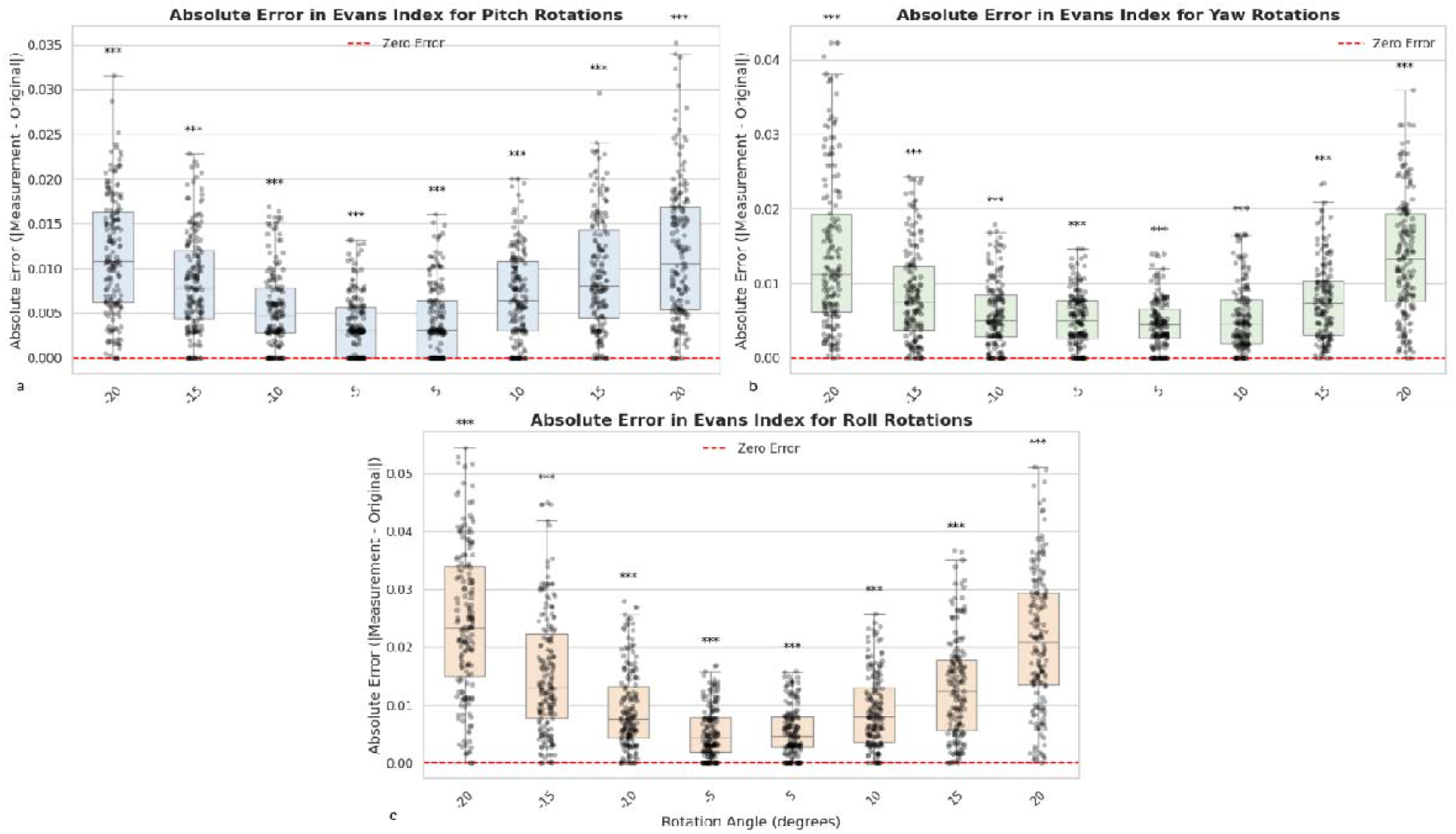
Effect of rotational misalignment on Evans Index error. Boxplots illustrate absolute Evans Index measurement error under rotational perturbations relative to the AC–PC line across the three orthogonal axes: (a) pitch, (b) yaw, and (c) roll. Each axis is systematically tested at eight rotation angles (±5°, ±10°, ±15°, ±20°). ^***^ indicates p-value < 0.0001

Furthermore, we examined the correlation between the ratio of lateral ventricle volume to intracranial volume (LV/ICV) and the Evans Index in both the NPH-only cohort (PENS) and the aging cohort (BLSA). As shown in Supplementary Figure 1, this analysis demonstrated a strong and statistically significant correlation in both cohorts (p < 0.0001 for each), with correlation coefficients of r = 0.68 in the NPH group and r = 0.84 in the aging cohort. (Supplementary Figure 3)

## Discussion

In this study, we developed and externally validated a fully automated deep learning–based pipeline for Evans Index (EI) calculation directly from raw T1-weighted MPRAGE MRI scans. Our method integrates landmark detection, automated AC–PC alignment, and robust segmentation of both the lateral ventricles and inner cranial vault, enabling accurate and reproducible EI measurement without manual intervention. Across internal and external validation datasets, including pre- and post-shunt NPH scans, the pipeline demonstrated excellent agreement with expert manual measurements, achieving minimal bias and high correlation coefficients. Importantly, the method remained robust across varying acquisition parameters and scanner vendors, and demonstrated sensitivity to subtle rotational misalignments, highlighting the importance of proper head positioning during image acquisition.

The clinical and research utility of automated EI measurement is substantial. EI remains a core morphometric metric in the diagnosis and monitoring of normal pressure hydrocephalus (NPH) and other disorders affecting cerebrospinal fluid (CSF) dynamics ^4,6,16^. Manual EI measurement, while straightforward, is prone to intra- and inter-observer variability, particularly in multi-center studies where slice selection and AC–PC alignment may differ ^8,17^. The proposed pipeline eliminates these sources of variability by leveraging volumetric data, standardizing orientation, and automating landmark and segmentation tasks. This level of reproducibility is critical for large-scale neuroimaging studies, where manual measurement is impractical, and for clinical workflows requiring rapid, objective assessment. The ability to process unprocessed T1 MPRAGE scans directly enhances applicability across both retrospective datasets and prospective clinical imaging.

Beyond accuracy, a key strength of this approach is scalability. By operating entirely on raw volumetric MRI data without the need for manual preprocessing, the pipeline can be integrated into high-throughput research environments, enabling the processing of thousands of scans with minimal human oversight. This is particularly valuable in population-based studies, such as the UK Biobank^18^, Alzheimer’s Disease Neuroimaging Initiative (ADNI)^19^, and the Baltimore Longitudinal Study of Aging (BLSA)^9^, where consistent and unbiased morphometric measures are required across diverse participants and imaging protocols. Moreover, automation democratizes access to EI measurement in clinical settings where neuroradiology expertise may be limited and supports telemedicine or cloud-based diagnostic workflows^20^. Such accessibility is especially relevant in under-resourced healthcare environments where NPH remains underdiagnosed or diagnosed late due to lack of standardized imaging metrics.

Based on the findings of this study, accurate alignment of MRI scans to the AC–PC plane is essential for precise Evans Index measurement. Previous automated approaches have largely relied on statistical registration techniques to approximate AC–PC alignment^21,22^. While these methods can approximate the target AC–PC plane, their indirect nature leaves them susceptible to alignment bias. Furthermore, the ventricular segmentation components in prior approaches have relied either on statistical or atlas-based methods, or on deep learning models trained on relatively small and homogeneous datasets^21,22^. Such limitations reduce their robustness, making them more prone to segmentation errors when applied to scans acquired with different imaging parameters or in the presence of structural abnormalities. In contrast, the proposed method directly addresses this limitation by integrating robust 3D landmark detection (BrainSignsNet) for explicit AC–PC alignment, coupled with deep learning–based segmentation (nnU-Net v2) trained on pathology-enriched diverse datasets.

This study has certain limitations. The external validation dataset, used to assess both ventricular segmentation and Evans Index calculation performance, consisted exclusively of patients with idiopathic normal pressure hydrocephalus (NPH) presenting with ventricular enlargement and no other intracranial abnormalities such as tumors, hemorrhages, or other abnormalities. While the proposed method is designed to operate across a wide range of ventricular sizes, its performance in cases with very small ventricles, or in the presence of coexisting abnormalities, remains untested. Further evaluation across imaging datasets that encompass a wider range of clinical presentations and pathological abnormalities is needed to determine this method’s generalizability.

In conclusion, we present a fully automated, deep learning–based method for Evans Index measurement from raw 3D T1-weighted MRI scans, capable of delivering accurate, reproducible, and orientation-standardized results. By eliminating manual intervention, the pipeline facilitates large-scale research, enhances accessibility in clinical settings, and supports standardized diagnosis and monitoring of NPH and related disorders. Future work will focus on expanding modality compatibility, optimizing robustness to extreme orientations, and integrating this tool into real-time clinical workflows.

### Data Sharing Statement

The complete codebase for Automated Evans Index Measurement pipeline, including all scripts and implementations of the BrainSignsNet, ICV and Ventricular Segmentation, and Evans Index Measurement code, is publicly available at https://github.com/SiavashShirzad/AutomatedEvansIndex The model weights used in this study will be provided upon reasonable request from the corresponding author and are planned to be released publicly in a future update.

## Acknowledgments

This research was supported in part by the Intramural Research Program of the National Institutes of Health (NIH). The contributions of the NIH authors are considered Works of the United States Government. The findings and conclusions presented in this paper are those of the authors and do not necessarily reflect the views of the NIH or the U.S. Department of Health and Human Services. Additional support was provided by the National Institute on Aging (Grant Numbers U19-AG033655 and P30-AG066507) and the National Institute of Neurological Disorders and Stroke (U01-NS12276). We thank the PENS Trial for their contribution of pre- and post-shunt surgery data, which served as the validation dataset for external validation in patients with Normal Pressure Hydrocephalus.

