## Supplementary Figure 1 for "Fully Automated Deep Learning-Based Pipeline for Evans Index Measurement from Raw 3D MRI"

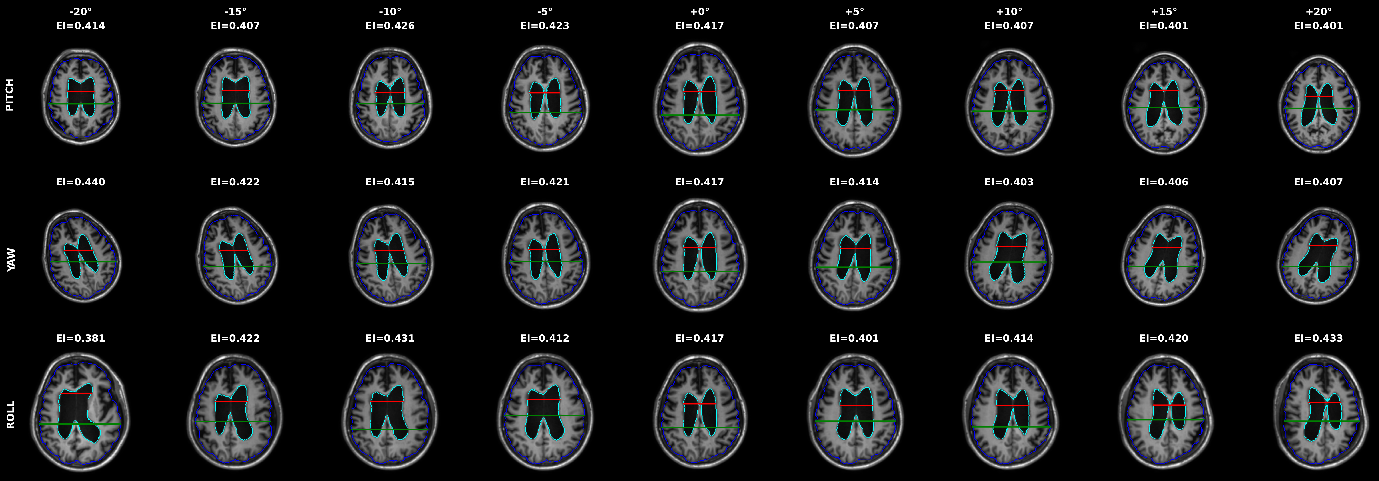

**Supplementary Figure 2.** **Visualization of MRI slices demonstrating rotational perturbations relative to the AC-PC line. Examples are shown for (a) pitch, (b) yaw, and (c) roll at selected rotation angles ( ±5°, ±10°, ±15°, ±20°) and the ground truth in the middle column. For each rotation, the corresponding Evans Index (EI) measurement is displayed above the image. The blue contour outlines the intracranial space, the cyan contour delineates the lateral ventricles, the red line marks the maximum width of the anterior horns of the lateral ventricles, and the green line indicates the maximum inner skull width.**
