## Supplementary Figure 2 for "Fully Automated Deep Learning-Based Pipeline for Evans Index Measurement from Raw 3D MRI"

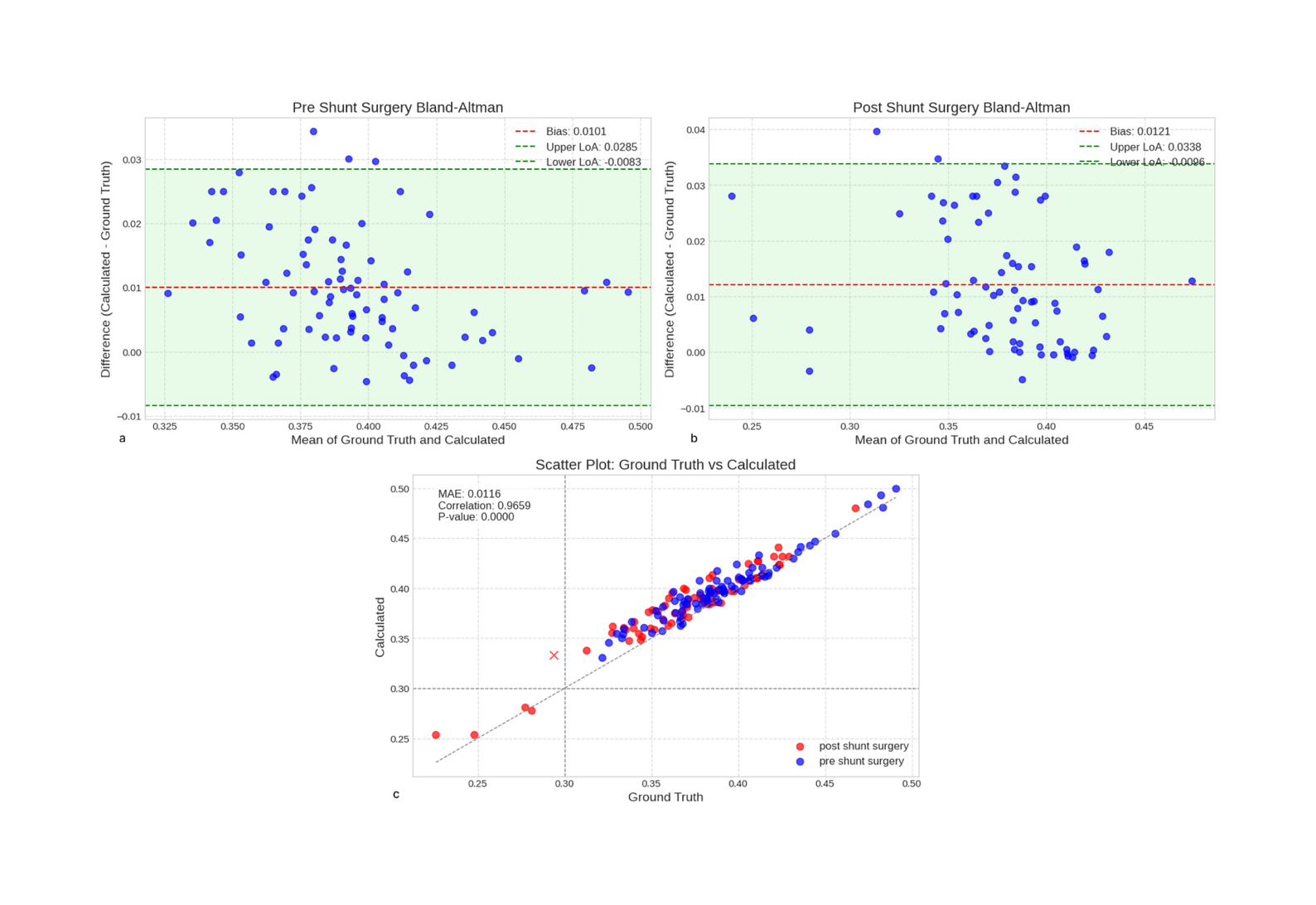

**Supplementary Figure 1.** Performance evaluation of automated Evans Index calculation using DLMUSE for lateral ventricle segmentation. (a–b) Bland–Altman plots comparing automated Evans Index measurements with expert annotations for pre-shunt (a) and post-shunt (b) scans. (c) Scatter plot showing correlation between automated and expert measurements, with pre-shunt scans in blue and post-shunt scans in red. Point(s) marked with X represent misclassified data at a 0.30 threshold.
