## Supplementary Figure 3 for "Fully Automated Deep Learning-Based Pipeline for Evans Index Measurement from Raw 3D MRI"

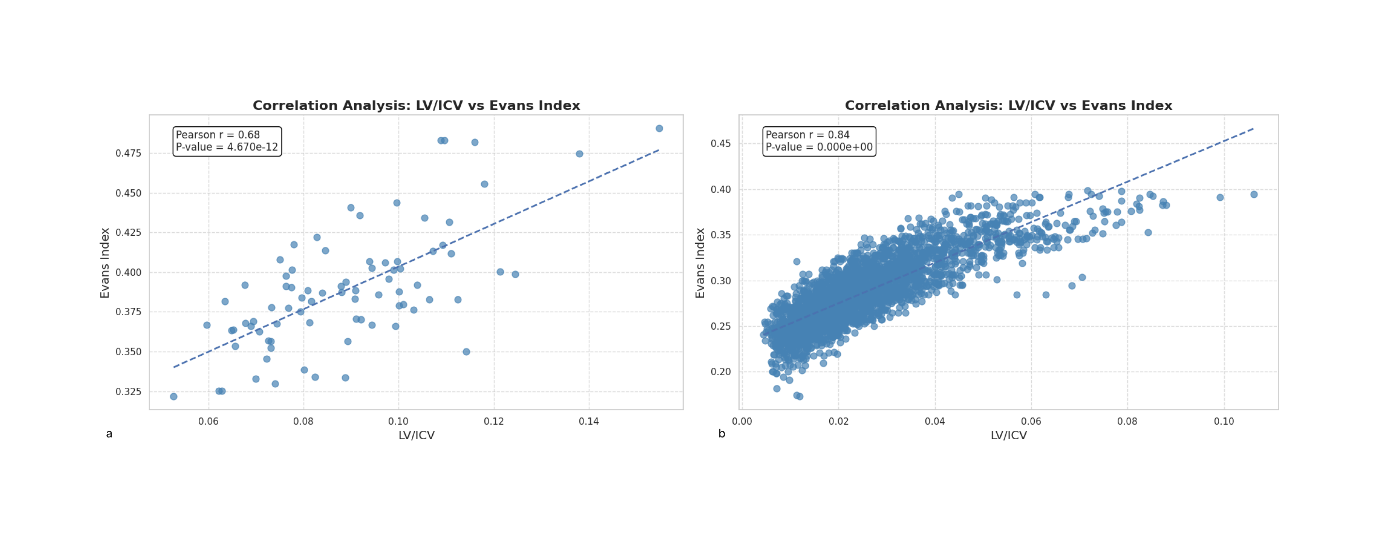

**Supplementary Figure 3.** Scatter plots illustrating the correlation between the ratio of lateral ventricle volume to intracranial volume (LV/ICV) and the Evans Index in (a) the NPH-only cohort (PENS) and (b) the aging cohort (BLSA). Both cohorts demonstrated strong positive correlations (P < 0.0001), with Pearson correlation coefficients of r = 0.68 in the NPH group and r = 0.84 in the aging cohort.
